# Testing the Feasibility, Acceptability and Effectiveness of the Problem Management Plus for Moms: Protocol of a Randomized Control Trial

**DOI:** 10.1101/2023.06.04.23290945

**Authors:** Irene Falgas-Bague, Maria Melero-Dominguez, Daniela de Vernisy-Romero, Thandiwe Tembo, Mpela Chembe, Theresa Lubozha, Ravi Paul, Doug Parkerson, Peter C. Rockers, Dorothy Sikazwe, Günther Fink

## Abstract

**Background:** Mental health disorders are one of the most common causes that limit the ability of mothers to care for themselves and their children. Recent data suggest high rates of distress among women in charge of young children in Zambia. Nevertheless, Zambia’s public healthcare offers very limited treatment for common mental health distress. To address this treatment gap, this study aims to test the feasibility, acceptability, and potential efficacy of a context-adapted psychosocial intervention.

**Methods:** A total of 270 mothers with mental health needs (defined as SRQ-20 scores above 7) will be randomly assigned with equal probability to the intervention or control group. The intervention group will receive a locally adapted version of the Problem-Management Plus and “Thinking positively” interventions developed by the World Health Organization (WHO) combined with specific parts of the Strong Minds-Strong Communities intervention. Trained and closely supervised wellbeing-community health workers will provide the psychosocial intervention. Mental health distress and attendance to the intervention will be assessed at enrollment and 6 months after the intervention. We will estimate the impact of the intervention on mental health distress using an intention-to-treat approach.

**Discussion:** We previously found that there is a large necessity for interventions that aim to address mother anxiety/depression problems. In this study, we will test the feasibility and efficacy of an innovative intervention, demonstrating that implementing these mental health treatments in low-income settings, such as Zambia, is viable with an adequate support system. If successful, larger studies will be needed to test the effectiveness of the intervention with increased precision.

**Trial registration:** This study is registered at clinicaltrials.gov as NCT05627206. https://www.clinicaltrials.gov/ct2/show/NCT05627206

## INTRODUCTION

Mental health disorders are a major global health issue and a significant cause of disability.^1^ Depression, and anxiety, also known as common mental health disorders (CMHD) are the most prevalent mental health disorders, with approximately 300 million cases worldwide in 2019, affecting women in greater proportion than men. ^2, 3^ CMHD result from a complex interaction of social, psychological, and biological factors, and their development is related to chronic stress exposure as well as to acute trauma experiences. Ever since the beginning of the COVID-19 pandemic, there has been an increasing concern about the negative effects of the pandemic on individuals’ mental health, with the greatest effect seen among women living in under-sourced contexts, such as low- and middle-income countries (LMICs). ^4^ Among women living in LMICs, mothers in charge of children under five years old are considered a key group to study and intervene in,^5^ as CMHD may limit not only their ability to care for themselves but may also subsequently impact their children’s development, contributing to the South-North poverty gap.^6-8^

The COVID-19 pandemic has increased the risk of developing CMHD among mothers living in sub-Saharan Africa.^5, 9^ In Zambia, where women are primarily responsible for caring for young children were already suffering from high rates of distress, the COVID-19 pandemic and its consequences in society (e.g. loss of jobs, closure of markets, reduction of healthcare visits) have exposed them to even higher levels of stress. Recent findings suggest that 26.1% of women in charge of young children suffer from mental health distress and that younger and less educated mothers were the most vulnerable during the COVID-19 pandemic, with higher mental health deterioration. ^5^ Despite the high rates of mental health needs, CMHD usually remain unnoticed and mainly untreated in this setting.^10^

Undetected and untreated mental health problems adversely affect women and their children in many ways, for example, by reducing well-being and quality of life, increasing food insecurity, and deteriorating mother-child relationships. Altogether, mothers’ mental health distress can result in inhibited child development and an increased risk of anxiety, reduced physical growth, cognitive impairment ^11^ and higher risks of non-adherence to HIV treatment for both mother and child.^12^

Zambia faces several challenges in addressing CMHDs within its primary care system. Shortage of mental health professionals, unmet training needs, and lack of evidence-based psychosocial interventions adapted to the local context are currently the most important obstacles.^10, 13, 14^ Effective treatments for mental health disorders are widely available in high-income countries but rarely applied in LMIC settings. In fact, the public healthcare system in Zambia, similar to other neighboring LMICs, offers very limited mental health services at primary healthcare and community level. The existing mental health workforce is concentrated in a few psychiatric institutions and high-level hospitals, which are permanently full and focus on attending people with severe mental illnesses. ^15^

Evidence suggests that culturally adapted psychosocial programs, delivered by trained paraprofessionals or community health workers, can effectively reduce CMHD burden. Community-based psychosocial programs increase mental health care access while providing treatment of CMHD within the community, reducing the work burden of the specialized mental health workforce while increasing workforce capacity.^16, 17^ However, even though some psychosocial interventions have proven effective in treating mental health symptoms,^18^ there are to our knowledge no culturally relevant treatments adapted to women in charge of young children living in low-income settings, which may reduce the engagement of these interventions as well as their impact.^19^ Targeted programs should consider the main obstacles that women face in engaging these interventions (i.e. domestic duties and lack of autonomy) and include specific topics that specifically are of women’s interest (i.e. self-care routines, parenting activities or/and empowerment strategies).

As such, we will conduct a randomized clinical trial testing the acceptability, feasibility, and effectiveness of a specifically culturally adapted “Problem-Management-Plus-For Moms” program with a sample of mothers residing in the Lusaka district in Zambia.

## METHODS/DESIGN

### Aims of the study

The overall objective of this project is to assess the feasibility, acceptability and impact of an adapted Problem-Management-Plus-For-Moms (PM+FM) on mothers’ mental health and their relationship with their children in a representative sample of urban Zambian community. This larger objective can be divided into three specific aims:

Specific Aim 1: Assess the mothers’ acceptability of the PM+FM intervention (measured as >50% of sessions attended, satisfaction measure and qualitative feedback).

Specific Aim 2: Assess the feasibility of the PM+FM intervention within an urban Zambian context (measured as ≥ 1 session attended and offered and participants’ and providers’ qualitative feedback).

Specific Aim 3: Assess the impact of the PM+FM intervention on mothers’ wellbeing through the improvement of their mental health status and their maternal-child relationship.

### Study design

This is a randomized clinical trial nested within a parent trial testing the impact of a child-focused intervention on children’s growth, the Zamcharts trial (NCT0512042). To assess eligibility for the PM+FM study, we used data from the 790 women enrolled in the parent trial and living in Lusaka.

### Recruitment of participants

#### Eligibility criteria

The eligibility criteria for this study is 1) being enrolled in the Lusaka subsample of the larger study and 2) having a high SRQ at baseline (SRQ>7).

All selected women from the intervention group will be contacted and reassessed with the Self-Reporting Questionnaire (SRQ-20) and the Hopkins Symptoms Checklist 25.

#### Exclusion criteria

Females with active suicidal ideation (determined by Paykel scale with scores 4 or/and 5), ^20^ severe substance use (ACOK-SUD >4),^21^ or mania or psychotic symptoms ^22^ will be excluded and referred to a partnering psychiatrist for further evaluation and treatment.

#### Recruitment and engagement procedures

Trained interviewers blinded by the treatment condition will assess women’s final eligibility, minimizing the risk of reporting or social desirability biases. Interviewers will explain the nature of the study to the participants and answer all questions prior to obtaining written informed consent.

We anticipate starting with the identification of cases, screening, and enrollment for this trial in October through December of 2022. A post-treatment assessment will be conducted among participating mothers approximately four months after the completion of the baseline survey (May 2023), immediately after completing the intervention. A second assessment will be conducted 12 months after baseline (June 2023).

We plan to focus on engaging participants in each of the research components of the trial (assessments and sessions) through systematic engagement protocol including regular calls to participants and home visits after two weeks of non-responsiveness.

### Randomization of participants

All eligible subjects will be randomized with probability of 0.5 to either treatment or control group. Randomization will be based on a random draw generated by the senior author (GF) using the Stata SE 16.0 statistical software package.

### Treatment of participants

#### Experimental intervention

Participants in the intervention group will receive the PM+FM intervention program delivered by wellbeing-community health workers (WCHWs). The intervention has been developed specifically to target mothers of young children living in sub-Saharan Africa. It combines elements from the “Problem-Management Plus” intervention and “Thinking positive” intervention developed by WHO with some selected components of the Strong Minds-Strong Communities psychosocial intervention.

To develop the PM+FM intervention, we followed the Castro-Barrera framework for cultural adaptation of evidence-based interventions. ^23^ Therefore, the intervention is culturally adapted to women’s local context of social and cultural values and norms. The final intervention, PM+FM, is a cognitive-behavioral therapy-based transdiagnostic psychosocial intervention aiming to improve mothers’ well-being, including depression and anxiety symptoms and mother-child relationships. It follows a collaborative and patient-centered approach where an individual’s needs are central, and contents of the intervention are adapted to each person.

The intervention will be provided mostly individually (sessions 1 and 10 can be delivered in groups), organized in ten to twelve 1-hour sessions, and provided on a weekly basis by a specifically trained and supervised WCHW in women’s homes and/or by phone. It will last a maximum of 4 months and will combine psychoeducation, problem-solving and cognitive restructuring techniques, motivational interviewing approaches, effective communication and mindfulness practices.

**Table 1.** Intervention summary of contents

| **EM-PM+ Intervention summary of contents** |  |
| --- | --- |
| Session 1 | Symptom's assessment. Introduction of the program and confidentiality. Cultural Formulation interview to understand women's needs. Psychoeducation about mental health needs (understanding adversity) and maternal-child positive relationship. Relaxing breathing. |
| Session 2 | Barriers to change, managing problems and stress management strategies. Introduction to mindfulness. |
| Session 3 | Symptoms assessment. Managing Problems (Problem-solving technique), "get going, keep doing" strategies (Behavioral activation). Managing Stress strategy and mindfulness practice. |
| Session 4 | Managing Problems: understanding unhelpful thoughts. Positive upbringing strategies. Stress management and mindfulness practice. |
| Session 5 | Symptom's assessment. Importance of support for positive upbringing and stress management. Strengthening social support. Mindfulness practice. |
| Session 6 | Effective communication skills, Mindfulness practice. |
| Session 7 | Symptoms assessment. Improving healthy habits for me and my child (nutrition, sleep, and substance use). Mindfulness practice. |
| Session 8 | Positive thinking and self-compassion practice. Mindfulness practice. |
| Session 9 | Review of the techniques. Staying Well and Looking to the future (Self-care plan). Mindfulness practice. |
| Session 10 | Review of a self-care plan. Social resources and referral. Mindfulness practice. Closure and Ending the programme. |

Women with severe mental health symptoms or with severe substance use symptoms will be referred to our collaborating team of psychiatrists and psychologists from the University Teaching Hospital and Chainama Hospital in Lusaka. Medication prescription and monitoring will be tracked by the research team. Pharmacological treatment (indicated by the psychiatrist) will be subsidized by the project, in case participants cannot afford it.

Regarding the training and supervision of the WCHWs, we will identify and train 4 to 6 people with at least a high school diploma, and previous experience in working within the community and interested in mental health topics. The training program entails 4 full days of instruction followed by 4 weeks of role-plays with a 2-hour weekly group supervision. Instructional training starts by addressing program objectives, ethics, emergency protocol procedures, psychoeducation about common mental health problems (Day 1) and basic components of psychotherapy (i.e., motivational interviewing, cultural formulation, mindfulness practice and problem-solving techniques) (Day 2). During the third and fourth day of instructional training, we will focus on the specific sessions’ content, special clinical situations (i.e., substance use involvement, domestic violence, and suicidal thoughts), and training related to their integration in the treatment site, including working at participants’ home, communication and referral procedures. It ends with a review of the emergency protocol procedures. Instructional trainings will be video recorded for further use in subsequent trainings. After completing roleplays with two example cases and receiving weekly supervision, WCHWs will be assigned with 10 to 15 cases each. All role-plays and the first two cases’ sessions will be audiotaped and heard by the supervisor. The supervision protocol includes weekly individual and group supervision throughout the training process (individual supervision) and trial (group supervision) to support the WCHWs and to provide feedback on the session tapes, share the fidelity forms and to resolve issues related to the delivery (e.g., severe cases and drifting). We plan to collect data on training needs and on the basic knowledge of mental health among the WCHWs at the moment of hiring and every 6 months, to evaluate mental health care delivery capacity. The supervisors will be local psychologist or psychiatrists specially trained by the PI of this project.

The proposed intervention does not pose any known risk to the women participating. Similarly, if women are not interested in being part of the intervention, they can simply inform the study staff that they are no longer interested.

#### Comparator

The comparator (control group) will receive no intervention.

### Primary and secondary endpoints

#### Primary endpoints

To explore the effectiveness of the intervention, our primary endpoint will be SRQ-20 ≤ 7, 12 months after baseline (approximately 3 months after completing the intervention).

Feasibility will be assessed by the percentage of participants completing 1 or more sessions of the intervention.

Acceptability will be evaluated using the percentage of participants completing the program (≥6 sessions completed) together with qualitative feedback from participants and providers.

#### Secondary endpoints

The secondary endpoints will be the mental health distress post-treatment (measured by HSCL-25 <1.50), psychological outcome profiles to measure women’s wellbeing (PSYCHLOPS), mother-child interactions measured by a selected measures from the World Bank Toolkit to address mother-child interactions and child early development and functionality through WHO-DAS2.

**Table 2.** Outcomes

| Outcomes | Measures Exploratory Efficacy Outcome |
| --- | --- |
| **Primary Outcome Measures for EM-PM+ assessed at enrollment, and 6 months (Intent-to-treat analysis)** |  |
| **SRQ-20** | 20 items measuring risk of common mental health problem. Used in the intent-to-treat analysis to explore effectiveness of the intervention using control group (12-months follow up data from parent study). Cut off of $>7$ will determine presence of mental health distress. |
| **Secondary Outcomes (pre- post- analysis)** |  |
| **HSCL-25** | 10 questions anxiety, 15 depression; ($\alpha = 0.90$). Used for the pre-post treatment analysis. |
| **WHO-DAS2** | Functionality |
| **World Bank's Toolkit and Inventory<sup>17</sup>** | Selected measures from the World Bank Toolkit to address mother-child interactions and child early development. |
| **Feasibility** | % of 1 or more sessions completed. Completion rates; ease/burden. Qualitative feedback from participants and providers. |
| **Acceptability** | 1) Participation rates; 2) Satisfaction scale; 3) Relevance of MEP-E. Qualitative feedback from participants and providers. |

#### Data management system

All data will be collected and stored electronically using the Survey Collect (SurveyCTO) software. Survey CTO is tool intended to facilitate mobile data collection services. It consists of an Android app that replaces paper forms used in survey-based data gathering. It supports a wide range of questions and answer types and can be utilized without network connectivity. All data collection will occur offline on password protected Android devices. Data connectivity will only be used to send data collected in the field. Data storage will be on a local secured computer in Zambia and on ServeyCTO’s secure servers.

#### Data security, access, archiving and back up

Project data is only accessible to authorized personnel who require data to fulfil their duties within the scope of the research project. On all the documents, participants will be only identified by a unique participant number. Data and databases will not be shared with the public and will only be available (fully anonymized) for verification purposes of authorities or scientific journals (as condition to publish results) – upon request only. All study data will be archived for a minimum of ten years after study completion.

### Statistical Analysis

First, we will begin with descriptive analyses of all data by group, examining balance between treatment and control groups. We will describe feasibility and acceptability measures at the participant level. Then, we will proceed with a pre-post intervention analysis in the treated sample only, using the Hopkins Symptoms Checklist as a main outcome, to allow for the comparison of mental health symptoms before and after receiving the intervention.

The main analysis of this trial will focus on calculating effect sizes and trends of effectiveness between the intervention group and the non-treated group from the parent trial using endline data from parent trial. We will use standard linear regression models to assess the impact of the intervention arm on women’s mental health. The primary outcome variable will be SRQ-20, used as a dichotomous variable with the cutoff at seven. Separate models will be estimated with and without baseline covariates. Our primary model will be estimated following an intent-to-treat approach, we will also estimate complementary per-protocol models restricting the intervention group to women completing the core components of the intervention (≥ 6 sessions).

Qualitative data will be collected through semi-structured interviews with participants during the treatment completion assessment and the focus groups with the WCHWs providing the intervention. Qualitative analysis will use a thematic approach to inform and complement quantitative results on feasibility, acceptability, and efficacy of the PM+FM intervention.

#### Interim analyses

No interim analysis is foreseen.

#### Deviation(s) from the original statistical plan

Any deviations from the original statistical plan will be reported in the Methods section of the trial paper.

#### Handling of data

Missing data will arise when a participant refuses to answer a question or complete part of the examination. Missing data may also arise if a participant drops out of the study. All participants are informed that their participation is completely voluntary and that they may refuse to answer any of the questions asked and can stop the evaluation at any time as well as discontinue participation at any time without any consequences. Missing data on covariates will be imputed using multiple imputations using chained equations. Missing outcome data due to sample attrition will not be imputed; however, we will conduct and report analyses examining the extent to which attrition differed across treatment conditions.

#### Sample size calculation

30% of the women enrolled in the parent trial displayed mild to severe symptoms of depression and/or anxiety (SRQ-20 ≥8). For this study, we expect an attrition rate of 13% based on previous studies. With an expected analytical sample size of 100 women per arm we are powered to detect an intent-to-treat difference of 20 percentage points in the prevalence of depression at endline with power 0.8, and 23 percentage point difference in depression with power 0.9.

#### Datasets to be analyzed, analysis populations

The main analysis will primarily rely on the data collected in the 12 month survey. Baseline variables will be used to 1) assess the extent to which balance was reached through the randomization and 2) create a set of control variables used in adjusted models.

#### Participant confidentiality

All records will be collected electronically and stored on a secure server. For the analysis, all personal information will be removed, and a unique Participant Number (e.g. 2738) will be assigned to each participant. The Investigators will keep a separate confidential enrollment log that matches identifying codes with the participants’ names and residencies, preferably in the Trial Master File.

## DISCUSSION

### Dissemination of results and publication policy

#### Dissemination to the scientific community; include lead in publications

Data analysis will lead to a series of synthesized reports and publications in peer-review scientific journals, for which all principles of data safety and protection are considered, and any results are presented in a fully anonymized manner.

#### Information of community and policymakers

This study will be conducted in close collaboration with the Zambian Ministry of Health (MoH). We will present the results of this trial to the mental health office of the MoH (in their quarterly meetings) and to an advisory board formed by a local team of clinicians and policymakers.

#### Ethics, audits and inspections

The study protocol has been approved by the ethical committees from Northern-Eastern region of Switzerland and the University of Zambia. The study documentation and the source data/documents are accessible to the ethics committees and auditors/inspectors at all times. All involved parties will keep participant data strictly confidential.

#### Availability of data and materials

De-identified data will be made publicly available after the publication of the trial.

#### Competing interests

The authors declare no competing interests.

#### Authors’ contributions

GF and IF designed the study. MC and TT provided input into the design and coordinated the selection of localities with the government. IFB, TT and MC coordinated local study implementation, MC and DVR ensured data quality. MC, IRB, TT and TL supported all logistical aspects of the project and coordinated all research activities. All authors provided input to this protocol and approved the final version.

## Data Availability

Deidentified research data will be made publicly available when the study is completed and published.

## Funding and role of funder

This project is sponsored by the Eckenstein Geigy Stiftung and Professorship. The funders had no role in study design, data collection and analysis, decision to publish, or preparation of the manuscript.

**Figure 1.**
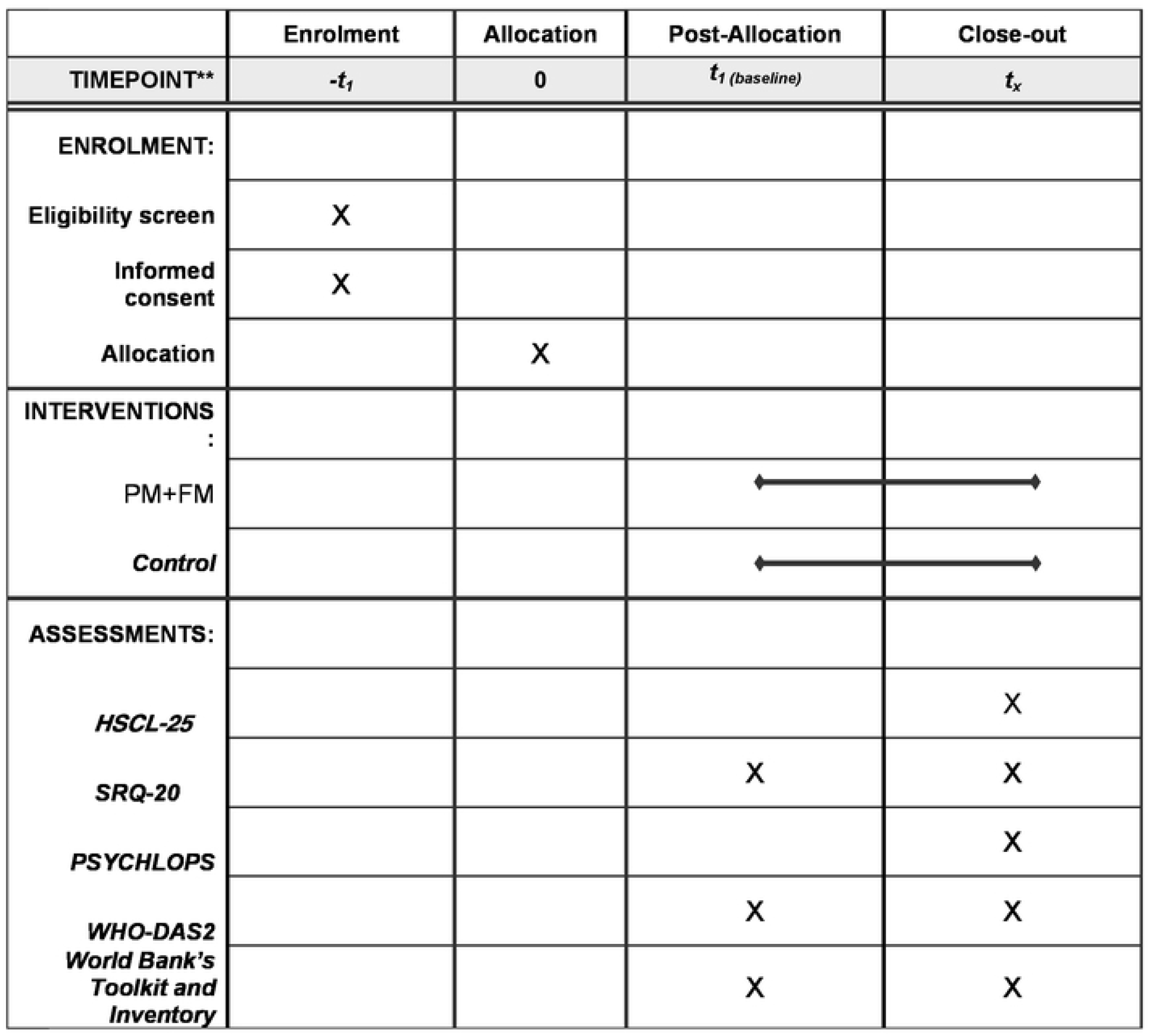

## Notes

### Competing Interest Statement

The authors have declared no competing interest.

### Clinical Trial

Registry: clinical trials.org Trial Number: NCT05627206. URL: https://www.clinicaltrials.gov/ct2/show/NCT05627206

### Clinical Protocols

https://www.clinicaltrials.gov/ct2/show/NCT05627206

### Funding Statement

The funders did not and will not have a role in study design, data collection and analysis, decision to publish, or preparation of the manuscript.

### Author Declarations

Ethics approval was obtained by the ethical committee of the University of Zambia

